# Stakeholder perspectives on the use of enhanced mobile phone capabilities for public health surveillance for non-communicable disease risk factors: A qualitative study

**DOI:** 10.64898/2026.04.22.26351443

**Authors:** Sylvia Nabukenya, Vicent Kasiita, Godfrey Bagenda, Joseph Ali, Elizeus Rutebemberwa, Dustin Gibson, Erisa Sabakaki Mwaka

**Author notes:** **Corresponding author:** Erisa S. Mwaka, Makerere University, College of Health Sciences PO BOX 7072, Kampala Uganda.

## Abstract

**Background:** Mobile phone–based tools are increasingly used to collect data on non-communicable disease (NCD) risk factors, particularly in low-resource settings where traditional data collection systems face operational and infrastructural constraints. This study examined stakeholder perspectives on the use of enhanced mobile phone–based capabilities to support the collection of public health surveillance data on NCD risk factors in low-resource settings.

**Methods:** An exploratory qualitative study was conducted between November 2022 and July 2023. Twenty in-depth interviews were conducted with public health specialists, ethicists, NCD researchers, health informaticians, and policy makers in Uganda. Thematic analysis was used to interpret the results.

**Results:** Four themes emerged from the data including benefits of using mobile phone capabilities for NCD risk factor data collection; ethical, legal and social implications; perceived challenges of using such mobile phone capabilities; and proposed solutions to improve the utility of phone-based capabilities in data collection on NCD risk factors. Participants recognized the potential of mobile technologies to improve data collection efficiency and expand access to hard-to-reach populations. However, concerns emerged regarding inadequate informed consent, risks to privacy and confidentiality, unclear data ownership, and vulnerabilities created by inconsistent enforcement of data protection laws. Social concerns included low digital literacy, unequal access to mobile devices, and fear of stigmatization. Participants emphasized the need for transparent communication, robust data governance, and community engagement.

**Conclusion:** Mobile phone–based systems can strengthen the collection of NCD risk factor data in low-resource settings; however, their benefits depend on addressing key ethical, legal, and social challenges. To ensure responsible deployment, digital health initiatives must prioritize participant autonomy, data protection, equity, and trust building. Integrating contextualized ethical, legal, and social considerations into design and policy frameworks will be essential to leveraging mobile technologies in ways that support inclusive and effective NCD prevention and control.

## Introduction

In 2021, non-communicable diseases (NCDs) accounted for 43.8 million deaths globally, with 73% occurring in low- and middle-income countries (1). Cardiovascular diseases accounted for at least 19 millions of these premature deaths, followed by cancers, chronic respiratory diseases, and diabetes, with the greatest burden increasingly concentrated in low-resource settings (1, 2). The effective prevention and control of NCDs depends on reliable, timely, and population-level data on behavioral and metabolic risk factors (3). However, traditional data collection systems in many low- and middle-income countries (LMICs) remain fragmented, resource-intensive, and limited in their ability to capture dynamic changes in NCD risk profiles (4).

There is a need for quality surveillance data to guide policymakers on how best to prevent and control the ever-increasing burden of NCDs. The World Health Organization recommends the development of innovative surveillance systems to strengthen the monitoring of NCDs and associated risk factors (5). The advent of artificial intelligence and advanced data collection techniques, combined with real-time analysis, has significantly impacted public health surveillance. The use of enhanced mobile phone capabilities - for example, location data, images, wearable mobile sensing devices, artificial intelligence technologies, and other innovative mobile healthcare applications - facilitates continuous health monitoring while providing timely alerts for potential health threats (6–9). As mobile phone ownership continues to expand across LMICs, mobile phone–based data collection tools are promising mechanisms to strengthen surveillance on NCD risk factors, expand geographic reach, and reduce operational costs (10, 11).

Mobile health (mHealth) platforms – including short message service (SMS) surveys, interactive voice response (IVR) systems, mobile apps, and passive data capture through sensors – offer significant potential for improving data quality and timeliness (12). These technologies enable remote participation, reduce the need for in-person data collectors, and allow individuals to report information in familiar and accessible formats. In contexts characterized by health worker shortages, limited access to health facilities, and logistical challenges, mobile phone-based data collection provides an alternative that can enhance inclusivity and support ongoing monitoring of NCD risk factors.

The integration of mobile technologies into public health data systems raises important ethical, legal, and social questions that have not been adequately explored. Mobile phones generate substantial amounts of sensitive personal data, including health information, location patterns, communication behavior, and other metadata that may disclose individuals’ private lifestyle attributes (13, 14). Limited digital literacy, low trust in institutions, and inadequate regulatory enforcement in many LMICs heighten concerns about privacy, data security, and the potential misuse of personal health information (15). Moreover, disparities in mobile phone ownership, gendered access to technology, and socioeconomic inequalities may result in exclusion or bias in NCD risk factor data (16).

Understanding the ethical, legal, and social implications (ELSI) of mobile phone–based data collection is crucial for guiding responsible design and implementation. Ethical considerations pertain to informed consent, autonomy, data protection, and prevention of harm (17, 18). Legal concerns include compliance with data protection laws, clarity on data ownership, and governance of public–private data partnerships. Social considerations include equity, trust, cultural norms, and the broader societal impact of digital data collection. Of note, several international and national laws and regulations govern data access, storage, and use of personal data with the goal of protecting individuals and communities from harm. For example, the European Union General Data Protection Regulation (GDPR) and the Organization for Economic Co-operation and Development (OECD) privacy guidelines emphasize transparency, user consent, data minimization, and strong safeguards to protect individual privacy across borders. In Uganda, the Data Protection and Privacy Act (DPPA) 2019 regulates the collection, storage, processing, and use of personal data, establishing obligations for data controllers/processors and rights for data subjects.

Despite the growing interest in digital health tools, there remains limited empirical evidence on how communities, end-users, and stakeholders in low-resource settings perceive and navigate these risks. Existing guidelines for digital health data management often assume robust regulatory frameworks and high digital literacy - conditions that do not always reflect realities in many LMIC contexts (19). As governments and health systems progressively adopt mobile technologies to address the rising NCD burden, a deeper understanding of these ELSI dimensions is urgently needed.

This study aimed to address this gap by examining stakeholder perspectives on the use of enhanced mobile phone–based capabilities - for example, location data, images, wearable mobile sensing devices, artificial intelligence technologies, and other innovative mobile healthcare applications – to support the collection of public health surveillance data on NCD risk factors in low-resource settings. By drawing on stakeholder experiences and contextual realities, this study provides guidance for responsible implementation of digital health solutions that are equitable, trustworthy, and consistent with the rights and expectations of the communities they aim to serve.

## Results

Twenty key informants (KI) were purposively selected to participate in the study, most of whom were male, researchers /academicians, and had a master’s degree (See Table 1).

**Table 1.**
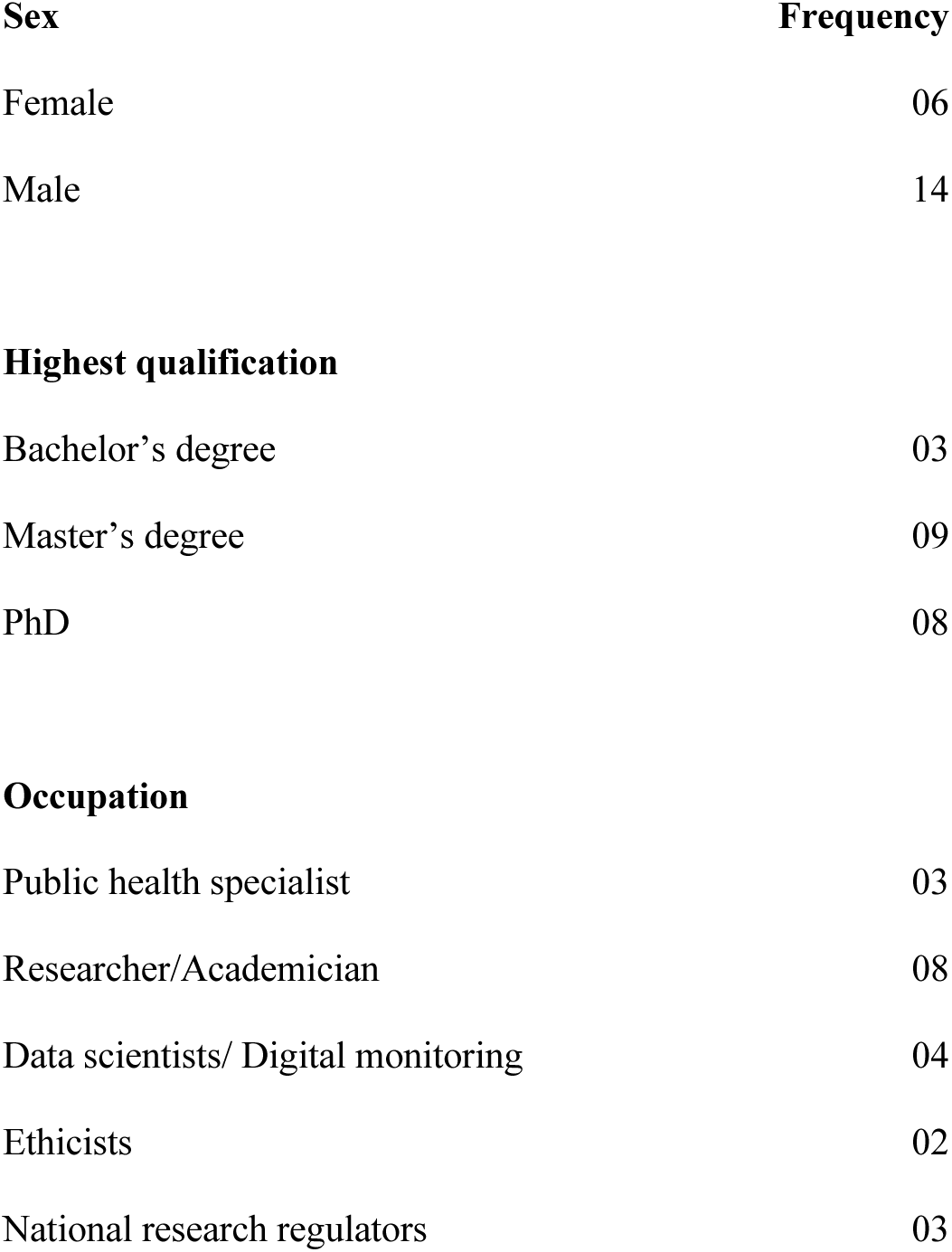
Key informant demographic characteristics.

Four themes were identified from the data:

i. Benefits of using enhanced mobile phone mobile capabilities for data collection on NCD risk factors
ii. Ethical, legal and social implications of collecting NCD risk-factor surveillance data using mobile phone capabilities.
iii. Perceived challenges of using enhanced mobile phone capabilities for data collection on NCD risk factors.
iv. Proposed solutions to improve the utility of mobile phone-based capabilities in data collection on NCD risk factors.

### Theme 1: Benefits of using enhanced mobile phone capabilities for data collection on NCD risk factors

All KIs considered the use of enhanced phone technologies beneficial with potential for improving NCD care and management. Data collected using mobile phone apps (such as a fitness tracking app for monitoring individual physical activities) were considered more accurate than asking individuals about their activity levels. These benefits were considered at both personal and health system level.

#### Benefits to researchers of using mobile phone capabilities

Enhanced mobile phone capabilities can be used to collect timely and accurate surveillance data. Of note, four informants surmised that people often may not remember their health history, leading to the collection of inaccurate data. However, they opined that using certain mobile phone applications, for example, wearable devices and location data, could help collect more accurate data than directly asking individuals.

> *Using phone advanced technologies enables the researcher to collect detailed data that a participant may not remember if asked. You cannot ask someone to mention places he went to in a week, and he is able to remember but an app will give you all that in time.* (KI 17)

Mobile phone technologies were also considered important in documenting people’s daily activities, follow up, early detection, and timely treatment of NCDs.

> *We* [researchers] *can actually be able to help the participant by knowing their behavior and what they are going through and we inform them early about the future harms. Let me give you an example, we have applications across sections in health. We have counselling sessions whereby someone answers a question and you are able to determine the level of anxiety, depression, stress. At the end of the day, the app goes ahead to show you what you are supposed to do. With this technology, a person can be informed about how they are behaving in order to improve their habits so that they do not get these diseases*. (KI 12)

> *…for example, if you are monitoring heart rate, mobile technology will enable you get to know what is happening at real time and then provide preventive information prior to any major challenges that may occur… So, in the end you are able to control or to prevent the worst.* (KI 05)

#### Benefits to individuals of using enhanced phone technologies

Key informants mentioned that enhanced phone technologies are non-invasive and use machine learning to predict a person’s susceptibility to NCDs such as diabetes and cancer.

> *So looking at the technological aspect, if we have what we normally call non-invasive technologies to monitor that […] and I think that is where we are having the aspect of machine learning coming in. Where you are trying to predict this person based on the behavior whether you might develop cancer or diabetes. Because some of these NCDs are more of behavioral aspect. (KI 06)*

These non-invasive predictive mobile applications were perceived as important in the early detection of NCDs. They were also considered essential in NCD prevention because they provide vital information on issues like caloric intake, weight loss, lifestyle modification, and healthy living. One KI stated that enhanced phone technologies can also be used for building social networks of people with shared lived experiences to learn from each other.

> *These technologies can be used to build a social network of people living with the same illness. For example, I know of apps, including WhatsApp, where people create safe spaces to share things that affect them and how they have overcome some health challenges. Participants tend to even share more on such platforms than when they are with researchers…* (KI 02)

### Theme 2: Ethical, legal, and social implications of collecting NCD risk-factor surveillance data using enhanced mobile phone capabilities

Despite the benefits of using enhanced mobile phone capabilities in collecting NCD risk factor surveillance data, informants raised several ethical, legal, and social concerns that may affect participants and their communities.

#### Ethical implications

Three main ethical issues were raised: informed consent, privacy and confidentiality, and trust. Six KIs raised concerns about the validity of consent when using certain mobile phone applications for public health surveillance. They doubted the voluntariness of consent because some mobile apps do not provide adequate information to guide users’ decision-making. Two KIs felt that consent should be ongoing and should involve regular sharing of information. However, this is not the case with many mobile phone apps.

> *There is that issue of consenting digitally though they are trying to see how best to use it. But consent should involve regular sharing of information and keep checking whether participants are still interested …. E-consenting should do this well…* (KI 09)

Further, two KIs contended that these mobile apps do not provide opt-in or opt-out options to end users.

> *But you find that even when you get these apps, the first thing they tell you is to accept before you can even use it. So, you just accept it because you want to use. However, it is not usually out of your own will…* (KI 01)

> *It will be safe if at least the developers to give options on how patients would like to be involved in such studies. Especially, also, how their data may be used.* (KI 15)

Another issue raised was the need for parental guidance since some mobile apps have age restrictions, yet it is difficult to verify the true age of user.

> *You find that anyone below 17 years is under age and cannot take sole responsibility as there will be need for parental consent. Some of these technologies can be harmful to children, especially teenagers who are too inquisitive and experimental.* (KI 10)

Several major privacy and confidentiality concerns were raised. Key informants indicated that people are usually worried about the safety of their information and frequently ask questions, as illustrated below.

> *People would be worried about privacy and confidentiality. They would worry about the end use of data, “who accesses the data?”, “how is it protected?” and those are some of the worries that are not here…* (KI 15)

Concerns were also raised about the robustness of the data security systems. The importance of ensuring that personally identifying information is secure and protected from unauthorized access was emphasized. Key informants further asserted that before any digital health technologies are used for public health purposes, they should have robust security systems to ensure that data are adequately protected.

> *How do we ensure that the patients’ data is safe? So before adopting digital health technologies for public health, we need to ensure that the patient’s data is safe, we need to address ethical issues, and the costing of the technologies.* KI 05

The success of using enhanced mobile phone capabilities for public health surveillance depends on people’s trust. Four KIs mentioned that people in communities are hesitant to use mobile devices due to privacy concerns.

> *Most of the participants think that it is encroaching on their privacy. There is a scenario where we enrolled a participant and explained to her everything about the study and particularly the phone. We gave her the watch and she went with it home. After three months, this participant had changed her attitude towards the watch. She told us that people in her family had some concerns about the watch, that it makes everyone in the study aware of where they stay and her husband was concerned about his privacy too since she was staying with him in the same house.* KI 02

One KI said that some people in the public feel that these devices are used for spying and tracking their private activities. This, he said, greatly affects the acceptability and use of these mobile phone capabilities for public health purposes.

> *… the patients or the general population might not also know what ethical issues might arise but they are cagey about using these kinds of technologies as they might think they are being tracked or being followed. They do not have their privacy. Most people will not try to use these digital health technologies because of that, but I think with time maybe we can embrace it.* (KI 09)

#### Legal implications

Four legal considerations were identified from the data: ignorance of data-related laws and regulations, poor law enforcement, data ownership, and data sharing. Seven KIs revealed that mobile phone-based data collection involves handling sensitive health information, which is protected under national and international data protection laws. However, four KIs were unaware of the laws and regulations governing digital and mobile phone usage in the country. Some were aware of Uganda’s data protection law but had not read it. Others were unsure of the government agencies that regulate the use of mobile phones and other digital gadgets.

> *I have not heard of any law that provides for security and privacy of my data, but if that is taken care of the fear is alleviated then digital technologies can be good to use.* (KI 03)

One key informant commented that there are various laws and policies governing digital information in the country; however, they are poorly implemented.

> *To be sincere the challenges are that we take long to implement what we put in place, so the laws that would be in place are not implemented. We keep reading about them but we never see them operating ….* (KI 07)

Key informants called for stricter enforcement of data-related regulations to ensure that personally identifiable information is adequately protected and not misused.

> *The guidelines should be clear on who owns the data and who should or shouldn’t use the data collected using anything to do with phones because phone use involves so many parties including handset owners, sim card owners, telecom companies, regulators etc* (KI 17)

It was noted that there are several players in mobile phone-driven public health surveillance, including the Ministry of Health, national regulators, the general public, mobile service providers, research and academic institutions, digital innovators, and others. Key informants called for stricter enforcement of data-related regulations to ensure that personally identifiable information is adequately protected and not misused, and that whoever breaks these laws should be penalized.

> *If they misuse people’s data, they too should be penalized…* (KI 013)

Another prominent issue was data ownership. Key informants felt that these mobile phone applications gather substantial personally identifiable information, some of which is collected without the app user’s knowledge. They wondered who really owns such data and whether the data providers have any say when such data are shared with third parties or used for innovations.

> *…also, there could be so much unknown information stored about you which you may not even know. This is because apps are able to pick so much information unknowingly and some researchers maybe do not even care or do not know that data is being captured and used by other people that participants did not consent to…*(KI_013)

One KI surmised that the data should be jointly owned by the data collectors and the data providers. One informant said that the data should belong to the health sector.

> *This information belongs to the health sector; if it is diseases, IT (Information Technology) is just a tool being used. My privacy is about talking to the doctor. ICT is just a media. Instead of me coming physically, I am using IT. It should be overseen by the health sector. If they are wondering who should come in regarding sharing this data, it should be the health sector.* (KI 14)

Another issue that arose was data sharing. As has already been mentioned, some KIs opined that many researchers are unaware of the data-related laws in Uganda, particularly regarding data sharing. One key informant highlighted the need to sensitize researchers on the regulatory requirements for data sharing, particularly in international collaborations where data are routinely shared.

#### Social implications

Several social concerns regarding the use of enhanced mobile phone capabilities for public health surveillance were raised. Of significant concern was the risk of harm to families. Several KIs mentioned that many spouses, particularly in rural areas, share mobile phones with each other; as such, there is a risk of inadvertent disclosure of sensitive personal health information. One KI cited an example of domestic violence that arose in a project where individuals were provided wearable devices for surveillance purposes.

> *There are some women whom we give the watches and their husbands want to know where they have gotten the watches from. I have heard about two scenarios where for one, the participant’s husband destroyed the charger and cut it into pieces while the other destroyed the watch completely…. The participants said that their husbands were feeling inferior. When the watch is off, we would call the participants to find out why we were not getting data from them. This made their husbands to start feeling insecure.* (KI 02)

Trust issues were also raised. It was noted that the community holds healthcare professionals in high esteem and trusts them. However, considerable suspicion is attached to programs that involve tracking people’s activities and movement. Skepticism and fear of the unknown were said to significantly affect the acceptability and full utility of digital technologies for public health. One KI pointed out that this lack of trust is usually compounded by societal misconceptions due to lack of community education and sensitization, as he mentioned a scenario where a participant said that the watch would expose her to cancer. He highlighted the importance of creating awareness and providing feedback to communities.

> *So well, we ourselves [researchers] the aspect of communicating back the findings either through different ways of dissemination that we have, but ensuring that the community where we collect this data from is made aware of the findings or what we found. I think that would go a long way in creating awareness in the community.* (KI 06)

The use of apps and wearable devices was also perceived to increase the risk of stigma.

> *This would affect confidentiality but also it can be stigmatizing. Many people do not want to be associated with any disease. So, if you want them to have some apps on their phones to collect NCD data, they will say that community will tag them as the sick which they do not want.* (KI 13)

### Theme 3: Perceived challenges of using mobile phone capabilities for data collection on NCD risk factors

Several challenges to using mobile phones for public health surveillance were identified. It was noted that many of these digital technologies are developed in the West and may be inappropriate for the local setting. Key informants pointed out that most applications are designed in English, making them inaccessible to a sizable proportion of the population because of low literacy levels in the community.

> *For these digital technologies most of our participants are illiterate and are unable to use smartphones. Regardless we try to make the apps easier for them but again there are those participants that will really fail.* (KI 05)

Full utilization of enhanced mobile phone technologies was also reported to be affected by the practice of sharing mobile phones among spouses. One KI opined that in such instances, there is a risk of collecting inaccurate data because the phone is used by an entire family.

> *You might find that there is only one smart phone in the family and because of the social cultural factors, it may be mainly owned by the man of the house and other family members cannot access it regularly… so whose information will be recorded or monitored? This may lead to inaccurate data.* (KI 14)

The increasing technology divide between urban and rural areas was identified as a major hindrance to using digital devices for public health purposes. Most KIs opined that good quality data can only be obtained using smartphones; however, they felt that these were expensive and unaffordable to the majority. Further, they indicated that the cost of internet was also unaffordable to many people.

> *…. the issue is social economic status as our population is very poor, so how do they access these devices? how do they afford internet data? It is not a matter of giving them the devices but you need to have people who are going to support these users to be able to make most use of these and this also requires resources; money.* (KI 08)

Another challenge identified was the difficulty in interpreting the data collected by some digital devices.

> *Some apps will tell you that you need to make 5000 steps in a day. However, what does 5000 steps equate to, what does it mean to the lay person? Is there a way of providing a simple explanation that 5000 steps help one burn a certain number of calories, which helps to maintain a good body weight and hence reduce chances of NCDs?* (KI 19)

### Theme 4: Proposed solutions to improve the utility of mobile phone-based capabilities in data collection on NCD risk factors

Several suggestions were offered on how to enhance the utility of mobile phone-based capabilities in data collection on NCD risk factors.

#### Meaningful community engagement and sensitization of the public about the use of mobile phones for public health purposes

Engaging communities is key to the success of health research and the introduction of health-related interventions. Eight KIs advocated for meaningful engagement of communities to ensure that people clearly understand these technologies, why the data are being collected, how these technologies are regulated, and the ethical implications of mobile health. They argued that this would improve acceptability and utilization for public health purposes.

> *I would advise them to ensure adequate community engagement, people should clearly understand why you would want to use their data. Participants should be made aware of the various laws and policies to protect them and they should as well know that everything is voluntary since people are always willing to participate when you are open. You also have to make sure that the regulatory processes are followed right from IRB approval, consent and administrative approval.* (IDI_018)

Low literacy was deemed a major challenge to effective utilization of enhanced mobile technologies for public health surveillance. Ten KIs recommended public education and sensitization about the different functionalities of smartphones and wearable devices to ensure that people in the community are informed about these capabilities. They also opined that mobile phone applications should be user-friendly, easy to navigate, and should have a provision for translation into the different local languages, with a guide on how to use them and how results can be interpreted.

> *I wish these apps can be public friendly where any lay man would be in a position to understand it very well. The technical people need to be creative to help with a preamble of what services the app can offer* (KI 06)

Three stakeholders advocated for introducing computer literacy in primary-level curricula. They argued that early exposure increases familiarity with such technologies and enhances their acceptability and utilization for public health purposes.

> *According to me, I think people need to just be educated. This should have started right from primary level… because they are already using these phones for other purposes such as sending money to others and therefore this should not be a problem. People need to be educated about: what information will be channeled through the digital technologies, for example, health related information, and how it will be kept private…* (KI 14)

#### Stakeholder engagement during the development of enhanced mobile phone applications

It was noted that most enhanced mobile phone technologies are developed by foreign innovators and simply imported into the country. As such, some of them are expensive, inappropriate, not well suited for the local setting, and unsustainable. Key informants recommended the engagement of various stakeholders during the development of these applications to ensure that they are affordable, context-specific, and easily adapted to local settings.

> *That is why it is good, that as you develop the tools, you need to involve the implementors, providers, the Ministry of Health people and others, so that whatever you develop as solutions is tailored to the needs of individuals and the country at large. Other than when you do very sophisticated trials of something that may not be applicable in a real-life situation. That is why some of these applications need implementation science research to understand “how do they fit in?”, “how do they address the barrier?” “What are the facilitators within the setting?”* (KI 14)

#### Creating awareness of data laws, regulations, and policies

Regarding the legal aspects, some KIs expressed the view that, while Uganda has many strong legal frameworks in place, their implementation and enforcement may be relatively weak, as KI 15 stated:

> *The guidelines are there to some extent, but I doubt if they are in practical implementation. Having a guideline and a policy is one thing but having it implemented is also another thing. The guidelines are there but their implementation is not there.*

The majority of KIs (15) recommended the education of researchers, public health specialists, the public, and other stakeholders on data protection and data-related laws, regulations, and policies.

> *There are laws and recommendations about these technologies. However, sensitization is key. Most of the health professionals do not know these laws. For example, you go to Ministry of ICT [Information and Communication Technology], Ministry of Health they will guide you on how to use digital health technologies. But awareness is minimal. I think we should address the awareness issue because I have tried to read. First, I used to think they were not there but there are some guidelines. But we need to sensitize and create awareness among the health professionals first.* (KI 05)

## Discussion

In this study, we sought to examine stakeholder perspectives on the use of enhanced mobile phone–based capabilities to support the collection of public health surveillance data on NCD risk factors in low-resource settings. As low- and middle-income nations increasingly use mobile health (mHealth) tools to improve data collection on NCD risk factors, it is critical to ensure that technology innovation is consistent with principles of autonomy, equity, justice, and transparency.

The KIs identified a range of benefits, including collection of timely and accurate information with minimal interference from recall bias, monitoring patients’ adherence to treatment, and timely sharing of information. However, our findings reveal tension between efficiency in data collection and the protection of participants’ autonomy. The use of enhanced mobile phone capabilities may compromise voluntariness, especially if the applications do not provide options for opt-in or opt-out, and may compromise the integrity of informed consent, especially when users have limited digital literacy. A recent study from Uganda reported that it is practically challenging to assess participants’ comprehension when utilizing mobile phone applications (18). Ensuring that consent remains voluntary, intelligible, and culturally appropriate requires provision of simplified information about the application (preferably in a local language), employing multimodal explanations (for example, audios or videos), and ongoing consent that enables participants to revisit their decisions over time (20). In this way, participants remain in full control over the use of their data for future research.

Data privacy and confidentiality raise significant concerns when using mobile phone data, whether collected actively through surveys or passively through monitoring sensors. A vast amount of sensitive information about an individual’s health behaviors, mobility, and social interactions can be collected by these devices, which, if not well protected, can lead to stigmatization of individuals, their families, and communities. Findings from our study raise concerns about unauthorized third-party access and data leakage, particularly in contexts where regulatory frameworks are not enforced. Strengthening data governance structures, clarifying data ownership, employing privacy-preserving technologies, and ensuring end-to-end encryption are crucial in building and sustaining trust in digital data collection systems (21–23). Moreover, stakeholders emphasized the significance of transparent communication about how participants’ data are stored, who has access, and what are the intended uses of their information are.

The legal and regulatory deficiencies also pose significant challenges. Although there are several existing regulations and policies in Uganda – for example, the Data Protection and Privacy Act, 2019, which requires lawful consent, transparency, and data minimization - enacting data protection legislation and the enforcement of such laws remain inconsistent. As stakeholders mentioned, many researchers using mobile phones for research and public health surveillance are not familiar with the data protection laws and regulations. Furthermore, mobile phone-based health research data often reside within regulatory ambiguities, particularly when generated by private sector entities operating outside established health systems (24). This raises concerns regarding cross-border data transfers, the commercialization of health data, and the lack of well-defined accountability mechanisms in cases of misuse (25). Our findings suggest that harmonized regulatory frameworks that are collaboratively developed with policymakers, communities, and technology developers are essential to ensure consistent safeguards while fostering innovation. In situations that require data transfer across borders, the investigators and research institutions should ensure that data sharing/transfer agreements are in place.

Stakeholders also mentioned several social implications arising from using enhanced mobile phone capabilities for data collection. While mobile phones can improve reach and inclusivity, there are several social factors that greatly impact their utility as public health tools. Socioeconomic status, digital illiteracy, cultural norms, the risk of stigma, and inequities in device ownership (especially among women) (26) all affect the use of mobile phones for disease surveillance. These factors may exacerbate existing disparities in the representation of NCD risk factors. Individuals without smartphones, stable internet access, or familiarity with digital platforms risk being excluded from research efforts, leading to biased datasets and potentially inequitable health policies. Addressing digital divides necessitates deliberate strategies such as providing alternative modes of participation, investing in digital literacy, and designing tools that are compatible with basic devices and offline formats (27, 28).

As mentioned by stakeholders, mistrust of digital technologies and fears of stigmatization related to certain NCD risk behaviors may influence participation and data accuracy. Engaging communities early in the design and implementation of mobile data collection systems can help build trust, improve cultural appropriateness, and increase adoption. Co-creation methodologies, such as participatory design workshops, active involvement of community advisory boards, and iterative feedback mechanisms, can ensure that tools are responsive to community values and expectations.

This study’s limitations include: first, stakeholders’ perspectives may not capture the full diversity of experiences across all demographic groups, particularly those without mobile phone access or with low digital literacy, who may face unique ethical and social challenges. Second, the rapidly evolving digital health landscape means that some legal and technological developments may have emerged after data collection, potentially limiting the contemporaneity of the findings. However, despite these limitations, the study provides valuable insights into the critical considerations necessary for the responsible use of mobile phone-based data collection on NCD risk factors.

## Conclusion

The increasing utilization of mobile phone-based capabilities for NCD risk factor data collection presents a significant opportunity to strengthen research data collection and facilitate timely interventions. However, this study highlights that the ethical, legal, and social implications of these technologies are substantial and require careful consideration. Protecting participant autonomy, safeguarding privacy, and addressing inequities are essential to building trust and ensuring responsible use of digital health tools. As nations progressively embrace mobile health technology, it will be essential to integrate robust governance frameworks, community engagement strategies, and equity-focused tools that ensure digital innovations support public health goals.

These findings highlight the need for closer scrutiny of the advance mobile health application used in collecting data for public health surveillance. There is also a need for research to determine which mHealth applications are appropriate and effective for public health surveillance in low resource settings where poverty, illiteracy, and other logistical challenges are profound.

## Materials and methods

### Study design and setting

This study was conducted between November 2022 and July 2023 and employed an exploratory qualitative approach (29–31). Key informant interviews were held with 20 officials and experts from central Uganda with knowledge of NCDs and/or related mobile health survey approaches. The stakeholders included officials from the Ministry of Information, Communication, and Technology; the Uganda National Council for Science and Technology (UNCST); research institutions; and Makerere University College of Health Sciences.

### Research team

The research team comprised bioethicists (SN, GB, JA and ESM), global health experts (DG, and ER), a social scientist with experience conducting qualitative research (VK), and two research assistants.

### Study procedure

Key informant (KI) interviews were conducted in English in convenient, private locations, mainly participants’ work offices. Interviews were scheduled at least two days in advance to ensure ample time for participants to prepare. Before conducting the interviews, the research team was trained on the protocol to ensure thorough understanding of the study objectives and procedures. Participants were contacted via email, which contained a brief description of the study and a request to schedule an interview. Expression of interest to participate was recorded through a positive response, after which a consent form was shared. Written informed consent was obtained from all participants, and they were assured of confidentiality. All interviews were conducted by SN, VK, and a research assistant. All interviews were audio-recorded with permission from participants, alongside detailed note-taking, and later transcribed verbatim. The interviews lasted between 30 and 45 minutes.

### Research tools

Data were collected using a semi-structured interview guide developed from the literature (32–34). The guide comprised open-ended questions on: 1) current and future use of supportive mobile phone technologies to conduct research and perform public health surveillance on NCD risk factors; 2) practical considerations; 3) perceived risks and benefits; 4) other values and interests; and 5) policy and oversight considerations. Open-ended questions facilitated natural discussion flow and provided participants a platform to share their thoughts, opinions, and experiences regarding the use of supportive mobile phone technologies for public health surveillance of NCD risk factors. The interview guides were first piloted with two researchers who were not included in the final study sample. Their feedback was used to refine probing techniques and rephrase questions in the interview guide. The research team held debriefing meetings after each interview to identify emerging perspectives not initially captured by the interview guide. Data collection continued until thematic saturation was achieved, when no new information or insights emerged.

#### Data analysis

Data were analyzed continuously throughout the study using thematic analysis (35, 36). All audio recordings were transcribed verbatim. Transcripts were verified for accuracy through word-by-word review while listening to the audio recordings to check for quality and spelling errors. Two authors (SN and VK) independently open-coded five transcripts. The transcripts were read line-by-line to generate the initial set of codes. Codes from the independent readings were synthesized through iterative discussion among the authors, and codes representing similar ideas were merged. Coding discrepancies between the two coders were resolved through consensus, after which a coding framework was developed. Final codes and themes were organized using NVivo 12 software (QSR International Pty Ltd., 2014). Themes were generated both deductively and inductively by considering predetermined and emergent themes (see interview guide). Three authors (SN, VK, and ESM) examined the themes for patterns consistency, achieving consensus on the final themes through several iterative discussions. All authors compared the emergent themes with existing literature to confirm that final themes accurately represented participants’ perspectives. Transcripts were also returned to three participants for member checking to verify whether the data collected accurately reflected their statements, thereby enhancing the credibility of study findings. The final codebook was iteratively refined to establish the themes presented in the Results section. We applied the Consolidated Criteria for Reporting Qualitative Research (COREQ) checklist to the presentation of findings (37).

## Data Availability

The data is freely available and is submitted as a supplementary file to this article.

## Ethical considerations

This study obtained ethics approval from the School of Public Health Research Higher Degrees and Ethics Committee (MUSPH-445) and Uganda National Council for Science and Technology (IS 128). The study was conducted in accordance with the provisions of the Declaration of Helsinki (38). Written informed consent was obtained from all participants prior to interview. All recordings and transcripts were de-identified, assigned special codes and stored on a password-protected computer. No participant-identifying information was collected.

## Financial Statement

This study was financially supported by the Bloomberg Philanthropies under the Data for Health Initiative (Grant No. 119668) awarded to DG and JA. The funders had no role in study design, data collection and analysis, decision to publish, or preparation of the manuscript.

## Acknowledgements

We acknowledge our research assistants who actively participated in participant mobilization and the data collection.

